# Increasing the Value of Digital Phenotyping Through Reducing Missingness: A Retrospective Analysis

**DOI:** 10.1101/2022.05.17.22275182

**Authors:** Danielle Currey, John Torous

**Affiliations:** Beth Israel Deaconess Medical Center, Harvard Medical School, Boston, MA, 02215, USA

**Keywords:** Digital Phenotyping, Mental Health, Smartphones, Precision Measurement

## Abstract

**Objectives:** Digital phenotyping methods present a scalable tool to realize the potential of personalized medicine. But underlying this potential is the need for digital phenotyping data to represent accurate and precise health measurements. This requires a focus on the data quality of digital phenotyping and assessing the nature of the smartphone data used to derive clinical and health-related features.

**Design:** Retrospective cohorts. Representing the largest combined dataset of smartphone digital phenotyping, we report on the impact of sampling frequency, active engagement with the app, phone type (Android vs Apple), gender, and study protocol features may have on missingness / data quality.

**Setting:** mindLAMP smartphone app digital phenotyping studies run at BIDMC between May 2019 and March 2022

**Participants:** 1178 people who partook in mindLAMP studies

**Main outcome measures:** Rates of missing digital phenotyping data.

**Results:** Missingness from sensors in digital phenotyping is related to active user engagement with the app. There are small but notable differences in missingness between phone models and genders. Datasets with high degrees of missingness can generate incorrect behavioral features that may lead to faulty clinical interpretations.

**Conclusions:** Digital phenotyping data quality is a moving target that requires ongoing technical and protocol efforts to minimize missingness. Adding run-in periods, education with hands-on support, and tools to easily monitor data coverage are all productive strategies studies can utilize today.

**Strengths and Limitations of this Study:**

○ Methods are informed by a large sample of participants in digital phenotyping studies.
○ Methods can be replicated by others given the open-source nature of the app and code.
○ Methods are informed by only mindLAMP studies from one team which is a limitation.

## 1 Introduction

The potential of digital technologies for deepening understanding of the etiology, risk factors, and course of diverse health conditions is now well known^1^. Research quantifying the lived experience of those with mental health conditions through capturing real-time information from a myriad of sensors is one of the fast-growing applications of these methods^2^. Specifically, utilizing digital phenotyping, the moment-by-moment quantification of the individual-level human phenotype with data from smartphones, it is possible to explore the cognitive, behavioral, and symptom domains of mental illness through smartphone data. Research has already emerged on digital phenotyping to understand the risk of suicide^3^, relapse in schizophrenia^4^, and severity of depression^5^. As this digital phenotyping research continues to expand, calls have emerged for a new focus on the resolution and psychometrics^6^ of these new digital measurements. Yet understanding the nature of digital phenotyping itself has proven challenging as this method is at times as dynamic as the illness it aims to quantify.

One under-researched and under-reported area in digital phenotyping work is the quantity of sensor data collection from smartphone devices. Sensors are often set to collect in cycles or at a certain frequency rate. We use the concept of data coverage to convey the expected quantity of data from a particular sensor as compared to the amount collected. For example, if the digital phenotyping app is set to collect accelerometer measurements at one reading per second (1 Hz), then there should be at least one data point collected in each one-second interval across that time, amounting to just over 1.2 million measurements collected in two weeks. Data coverage is rarely, if ever, 100%. Sensor non-collection can occur for multiple reasons including participants turning off data permissions, setting them incorrectly, or phone operating systems turning off background data collection for performance/battery reasons. Another challenge is the fact that the Android and iOS permissions required to collect sensor data evolve over time, presenting a moving target. For example, new rules around the low-power mode settings of Android and Apple smartphones mean that if the phone is ever in this mode, then sensor data permissions are likely temporarily revoked. If participants do not actively engage with the app (ie open it and take a survey), then permissions will likely also be revoked over time. Given these phone operating system details are proprietary, determining the source of missingness, and obtaining continuous, high data coverage can be difficult. However, it remains important to understand the level of data coverage that should be expected, the level needed for analysis, and to seek ways to increase data coverage towards this acceptable threshold. In this work, we sought to investigate data coverage in a combined data set of over 1000 participants collected using the mindLAMP app^7^ from 2019 to 2022.

Prior work has explored passive data collection by the Beiwe platform^8,9^. Kiang et al. at accelerometer and GPS data coverage from 211 participants from 6 different studies between 2015 and 2018 [8] and Staples et al. evaluated data from 16 participants^9^. Kiang et al. reported missingness of 19% for accelerometer and 27% for GPS^8^, which is impressive but may not be possible today in 2022 as sensor data permissions have changed in the four years since data was collected for that paper. Except for a slight difference in accelerometer between Black and White participants, the researchers did not find significant differences in missingness across race, gender, or education background^8^, but they did report a difference in GPS data coverage between Android and iOS users. Another large study with 623 participants using the RADAR platform examining data collected from November 2017-June 2020 found that only 110 participants had >50% data across all data types collected^10^. In addition, the researchers found no link between baseline depression and data availability in their sample, which suggests digital phenotyping methods are applicable regardless of illness severity. The authors concluded more investigation around this data quality is warranted before a more complex analysis is feasible. Thus, these studies highlight the need to further explore data coverage in larger samples and understand what is possible today for digital phenotyping methods with data collected today.

Obtaining high levels of data coverage is important because if data collection is sparse then derived features may be inaccurate. These challenges in digital phenotyping have parallels in other fields. For example, it is well known that participant motion in neuroimaging or eye blinks in electroencephalograms can introduce artifacts. Research protocols have evolved to minimize these sources of error both during data collection and during data processing. Likewise, such approaches are necessary for digital phenotyping and even more critical given data is collected outside of a lab or clinic, subject to changing availability from smartphone manufacturers, and related to on ongoing participant active engagement.

Beyond the impact on features, different data coverage standards and lack of reporting in the literature makes studies difficult to reproduce. Heterogeneity in digital phenotyping studies is a problem, for example, a 2022 review of digital phenotyping papers around depression reported such a high degree of heterogeneity that precluded any quantitative analysis^11^. In this same review, the authors found studies with both positive and negative correlations with digital phenotyping derived sleep measures and mood – underscoring challenges in working with this data in a mental health context. To understand the clinical implications of digital phenotyping data, it is thus critical to ensure any results are not related to missingness completely at random (ie smartphone app error), or missingness at random (ie different patients use the app differently). Although not the focus of this paper, determining the type of missingness in the raw data of digital phenotyping studies remains an important, multifaced challenge, and methods for addressing missingness have been proposed^12^. Here we focus on methods to reduce missingness in data and limit it to below acceptable thresholds.

This paper has three goals: 1) to present results on the coverage and missingness of passive data collected by the mindLAMP platform^7^ on a large data set, 2) to explore how poor data coverage can affect features and analysis, and 3) to introduce solutions to maximize the use of digital phenotyping data through increasing coverage of raw data sensor collection.

## 2 Method

### 2.1 Data Sets

Appreciating that there are many digital phenotyping platforms, our goal is not to suggest any single one is superior. While our examples derive from the open source mindLAMP created by our team at Beth Israel Deaconess Medical Center^7^, the principles discussed are applicable to any digital phenotyping app that collects sensor data and to any illness or condition being studied.

Patient involvement with the development of mindLAMP is reflected in the co-development of the app^6^ with all stakeholders before any research was conducted. All mindLAMP studies return data immediately to participants within the app. No patients were involved in setting the research question, outcome measures, or design of this study. Patients were not involved in the interpretation or write-up of the results. The results of this work will be shared with patient groups at the National Alliance of Mental Illness (NAMI) annual meeting.

We examined data from seven studies conducted by our team in the last two years. Each study received IRB permission and all participants signed written IRB informed consent. While the studies differed in their clinical populations (patients with schizophrenia; college students with stress, anxiety, and depression; patients with depression using mindLAMP to augment therapy), they all collected two common digital phenotyping data streams: geolocation and accelerometer data. Each study presented participants with a symptom survey at least every three days^13-16^. For completeness, all participants in each study (including those that were discontinued because of non-adherence to protocol) were included in this analysis. Study details can be found in Table 1. The number of participants in each study is listed in Table 2.

**Table 1.** Study details.

| Study | Enrollment criteria | Timeframe | Length of study |
| --- | --- | --- | --- |
| Study 1 (schizophrenia) | Referred by Clinician | May 2019-Nov 2021 | 52 weeks |
| Study 2 (college) | PSS ≥ 14 | Nov. 2020-May 2021 | 4 weeks |
| Study 3 (therapy) | PHQ-9$>$10 | Sept-March 2022 | 8 weeks |
| Study 4 (anxiety) | GAD-7 $>$ 5 | Oct-March 2022 | 6 weeks |
| Study 5 (schizophrenia) | Referred by Clinician | Aug 2021-current | 52 weeks |
| Study 6 (schizophrenia) | Referred by Clinician | Oct 2021-current | 12 weeks |
| Study 7 (college) | PSS $\geq 14$ | Nov.-Dec.<br>2021 | 4 weeks |

**Table 2.** Number of participants in each study. Discontinued participants were included in this analysis.

| Study | Total | GPS |  | Accelerometer |  |
| --- | --- | --- | --- | --- | --- |
| | | Has data | Coverage $> 0.5$ | Has data | Coverage $> 0.5$ |
| Study 1 (schizophrenia) | 92 | 54 | 17 | 56 | 13 |
| Study 2 (college) | 644 | 557 | 243 | 571 | 163 |
| Study 3 (therapy) | 92 | 88 | 73 | 87 | 75 |
| Study 4 (anxiety) | 220 | 198 | 146 | 202 | 151 |
| Study 5 (schizophrenia) | 22 | 15 | 13 | 15 | 12 |
| Study 6 (schizophrenia) | 12 | 12 | 10 | 12 | 11 |
| Study 7 (college) | 96 | 96 | 84 | 96 | 88 |
| Total | 1178 | 1020 | 577 | 1039 | 513 |

### 2.2 Passive data coverage

We define data coverage as the percent of expected data that was collected. For example, if we expected data to be collected every hour, we would check whether there is at least one data point in every hour of every day. If 23 of the hours had data, the coverage would be 23/24 = 0.958 for that day. In this work, we focus on accelerometer and GPS data. In mindLAMP, the data can be collected continuously at a preset maximum sampling rate. GPS is collected at a maximum sampling rate of 1 Hz and accelerometer is sampled at a maximum rate of 5 Hz. Of course, for various reasons data is often not captured at these maximum levels. However, to capture the health-related features that we are interested in, it was not necessary to have data sampled at the maximum frequency. For GPS we aim to assess the number of significant locations visited each day, so the data sampled must represent each new location. Based on the down-sampling experiment described below, we were able to obtain accurate information with GPS readings at approximately every 10 minutes instead of the maximum of one per second. For accelerometer, we are interested in computing jerk, the rate of acceleration changes over time, to identify active and inactive periods. Thus, we chose a 1 Hz sampling rate to explore data coverage as based on our experiences, this is the highest that most phones can reasonably achieve over a period of weeks and months.

We plotted this data to examine data coverage for each of our studies for GPS and accelerometer and how data coverage has changed over time. In addition, we calculated the number of days since an activity (ie survey, mindfulness activity, tip) was completed to investigate the relationship between active engagement with the app and passive data coverage.

### 2.3 Down sampling experiment

To simulate how decreased raw data coverage affects the quality of features derived from this data, we selected the participant at 98% 10-minute GPS data coverage with the highest 1-second data coverage (83%) (see Appendix A) and iteratively down-sampled their data. For example, to generate the 2s data we randomly selected one data point in each two-second interval. The intervals used simulated data collection every 1s, 5s, 10s, 15s, 30s, 60s, 5min, 10min, 30min, 60min, and 4hrs. We then computed home time and entropy on this down-sampled GPS data and found the difference between the features derived from the original full data.

### 2.4 Correlations with passive data

To further illustrate how passive data coverage can affect clinical results, we have presented correlations between passive data features from a study with college students. We calculated correlations between features derived from passive data for all days and for all days where GPS data coverage was above a 50% threshold. As an example, geolocation passive data can be used to assess home time and this feature can be correlated against screen duration. Our implementation of these passive data features and data coverage is open source and can be found at https://github.com/BIDMCDigitalPsychiatry/LAMP-cortex.

## 3 Results

### 3.1 Data coverage

Realizing that data coverage will continue to evolve with smartphone technology, we created violin plots to show the distribution of data across our studies which is shown in Fig. 1. Studies 1 and 2 were completed before Fall 2021 and have lower data coverage than more recent studies running on newer versions of the app (p < 0.001). As shown in the figure, coverage for accelerometer and GPS is similar. The trajectory of average data coverage across all participants in our studies over time is shown in Fig. 2.

**Fig. 1.**
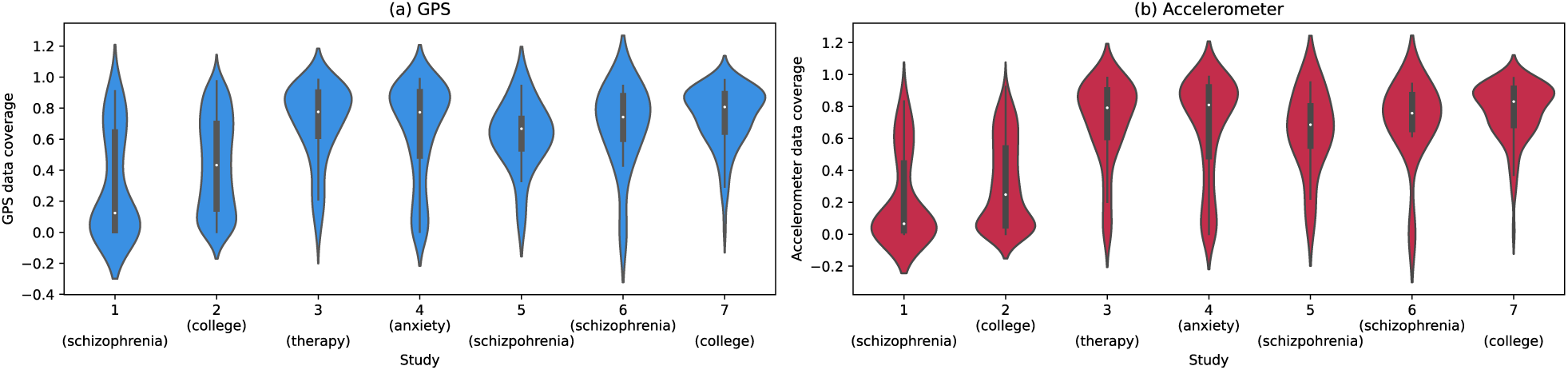
Violin plots of (a) GPS and (b) accelerometer data coverage for each of the seven studies.

**Fig. 2.**
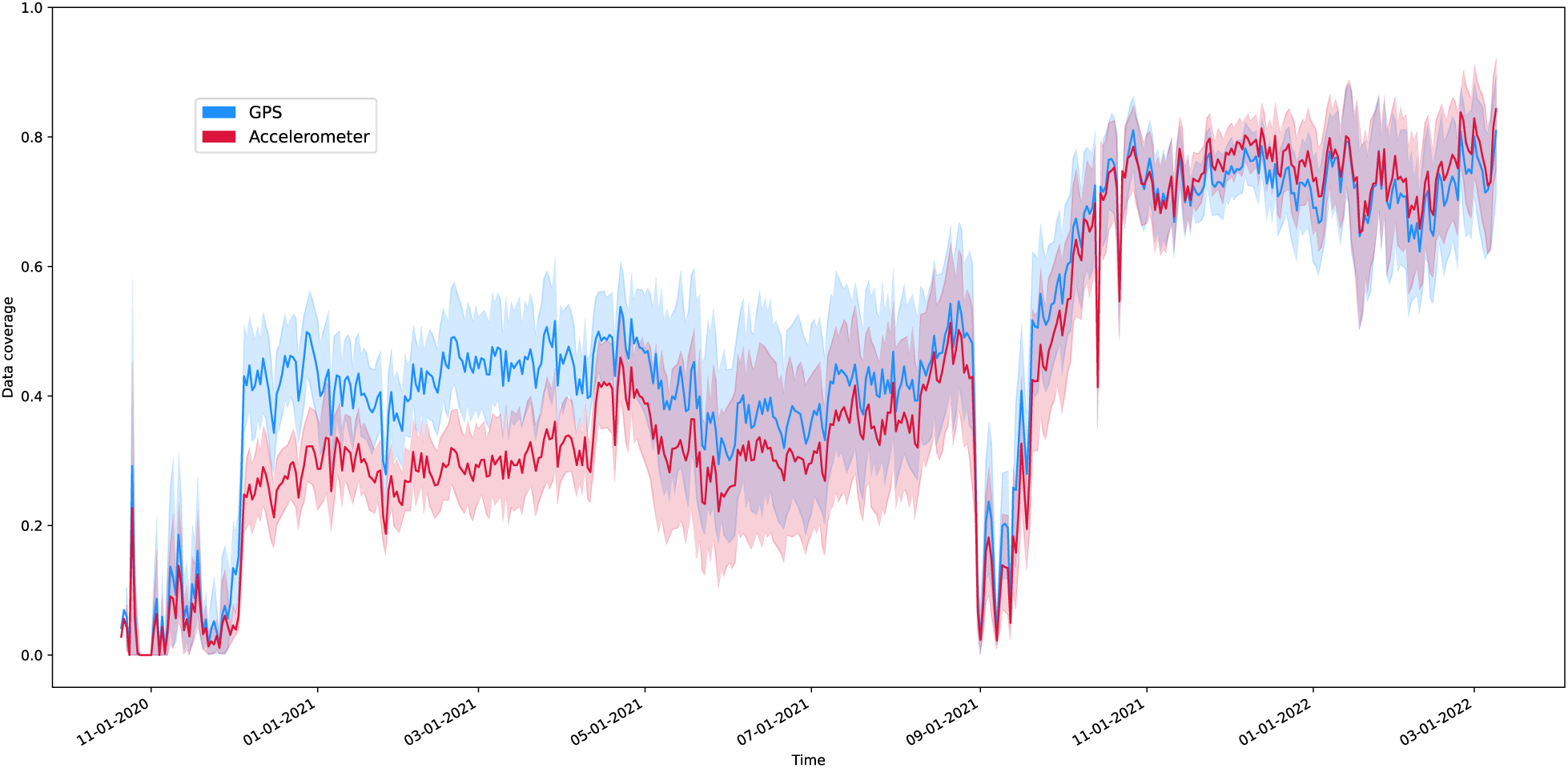
Mean data coverage across the studies over time. GPS is shown in blue and accelerometer in red. The shading indicates the 95% confidence interval for the line.

Within any single study, one reason that data coverage may be lower is that after the app has not been used for a certain period the phone’s operating system will halt data collection. This is likely related to battery-saving features enforced by the operating systems of smartphones. When the user completes any activity in the app (eg a survey, mindfulness activity, or even viewing their results), this active engagement likely delays the operating system from closing the app. The exact amount of time that must elapse before the operating system shuts down data collection is not reported by phone manufacturers and differs between phone versions but we can investigate it experimentally. To explore this effect, we plotted data coverage by the number of days since any activity was last completed. These curves are shown in Fig. 3 and appear different for Android and iOS, but both phone types show a downward trajectory with higher variance in data coverage after multiple days of user non-engagement. We have split the data by the older (Study 1 and Study 2) and newer studies in Appendix B. In both research and clinical terms, this finding highlights the importance of active engagement necessary to collect passive data with maximum data coverage.

**Fig. 3.**
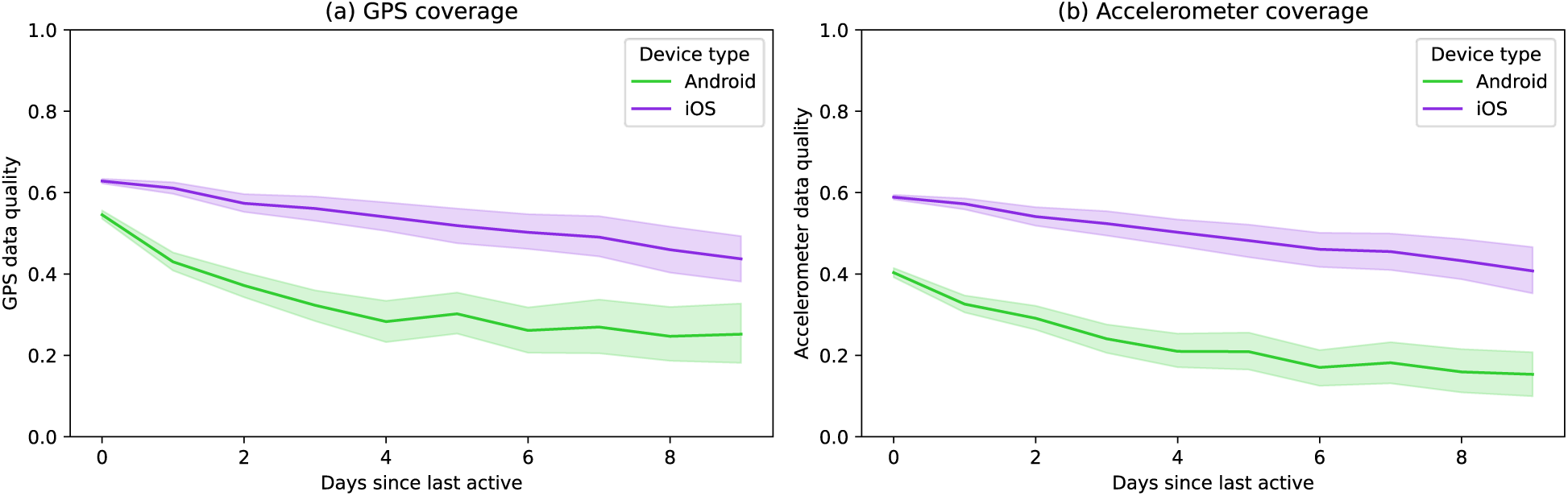
Plot of the (a) GPS and (b) accelerometer data coverage versus the number of days since the participant last interacted with the app. The Seaborn plotting library was used which generated a shaded 95% confidence interval for the data [18].

Finally, from Tukey’s HSD We found a significant difference in data coverage between male and female participants (p < 0.05) and a significant difference between African American and white, Latinx and Asian participants (p < 0.01). Moreover, we found a significant correlation between anxiety and GPS / accelerometer data coverage (p < 0.01) and between depression and accelerometer data coverage (p < 0.05).

### 3.2 Poor coverage and derived features

While passive data can provide interesting insights into a patient’s mental health, if coverage is low then the features derived from this data may not be accurate and could lead to false interpretations. Fig. 4 depicts an illustrative example of this problem using home time. Home time is computed by counting the GPS data points at each location visited by a participant and then finding the amount of time at the location with the greatest number of data points. If GPS sampling frequency is high enough to capture all the locations that a participant visits that day, we will be able to accurately determine the home location as shown in Fig. 4a. But if the GPS sampling frequency is low, as in the example in Fig 4b, then data may not evenly sample visited locations. In this example the “gym” location was sampled at a higher rate than “home”, so the incorrect home location was chosen, and home time was greatly underestimated.

**Fig. 4.**
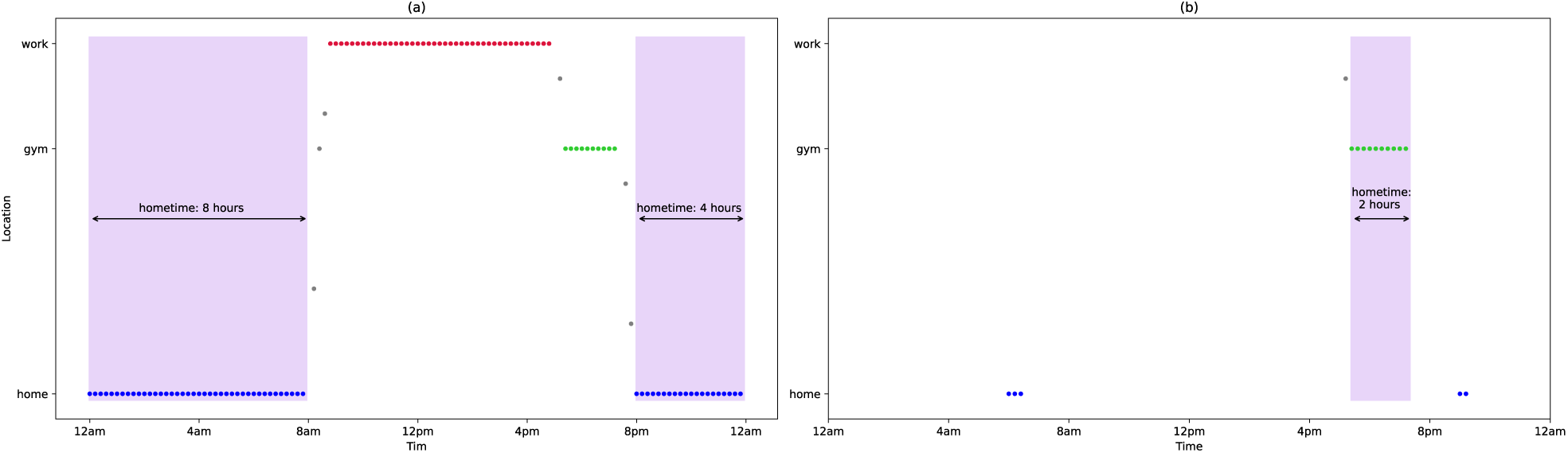
A simualted example of how low data coverage can result in inaccurate features. (a) shows GPS data collected consistently over time, resulting in accurate identification of the home location (blue dots) and correct duration at home (shaded in purple). (b) shows an example where GPS is not recorded consistently, and where the “gym” location has the most data collected and is incorrectly identified as the home location.

To illustrate this problem with participant data, we performed an experiment where we down sampled GPS data to try to mimic this data missingness. Fig. 5 shows how error in the computed home time and entropy begins to increase when points are only sampled every 5-10 minutes or less.

**Fig. 5.**
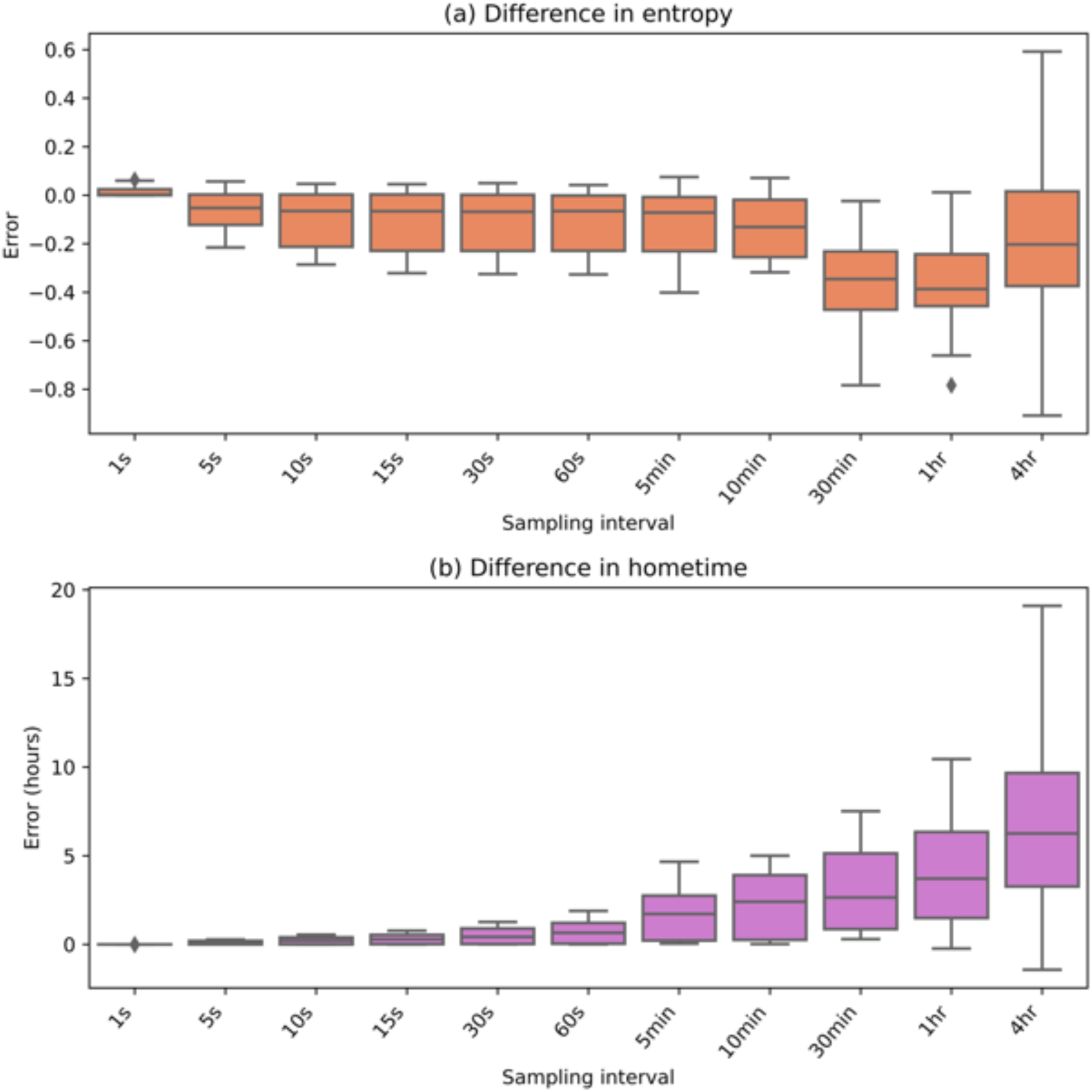
Error from the original feature values when raw GPS data is iteratively downsampled. (a) shows absolute error for entropy and (b) shows error for home time in hours.

When features are inaccurate, this will impact analysis results and clinical interpretations. To highlight this, we produced correlations between passive data derived features from one of our studies with college students. We used either all days of passive data regardless of data coverage (Fig. 6a) or only included days with GPS coverage of greater than 0.5 (Fig. 6b). 321 of the 2206 total days of data (14.6%) of the days had GPS coverage of less than 0.5. We can see that the correlations change depending on which passive data is used.

**Fig. 6.**
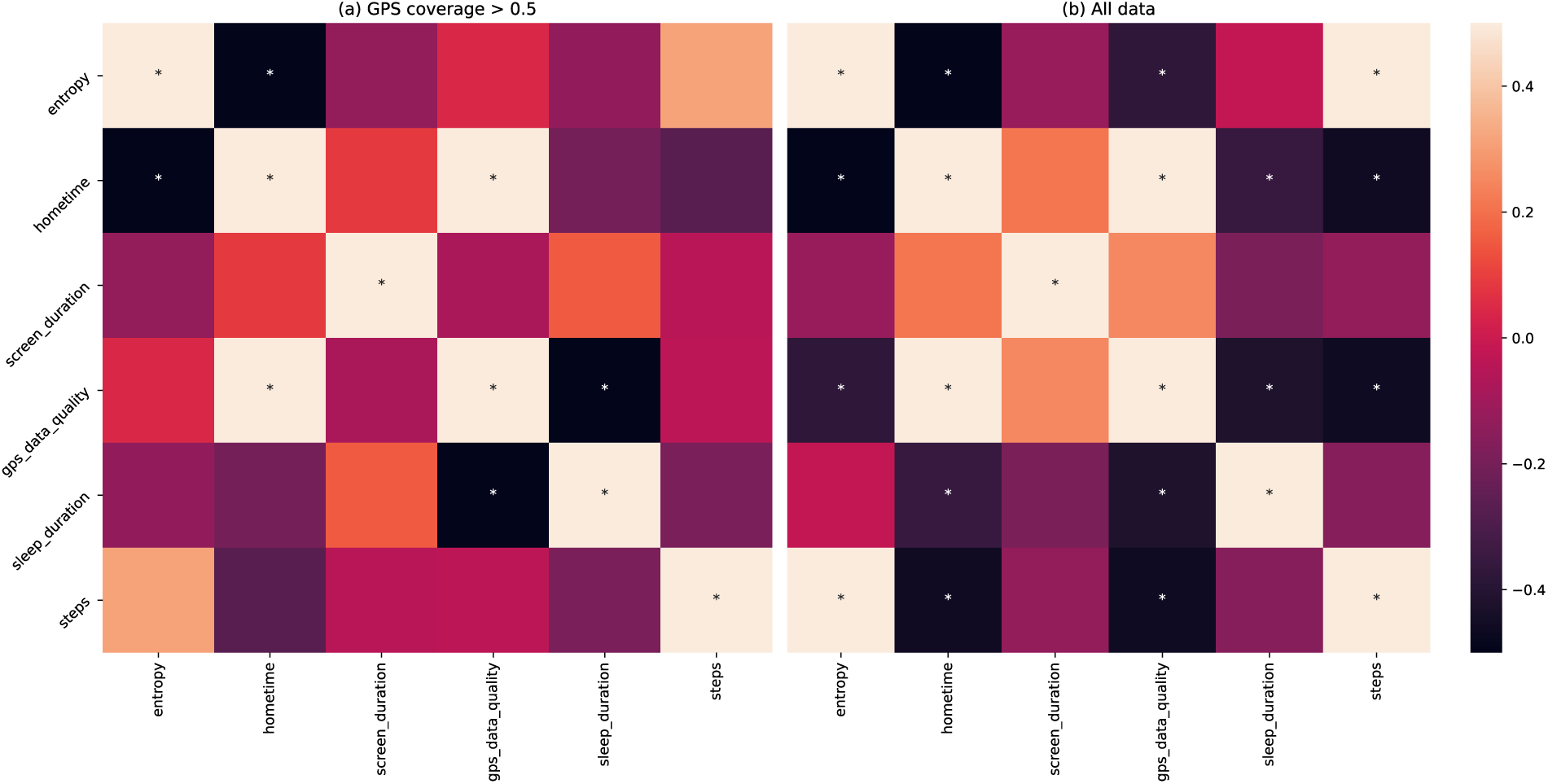
Correlations between the average value of different passive data features using (a) only days of data where GPS coverage is greater than 50% and (b) all data. Significant correlations (p < 0.05) are marked with a *.

### 4 Suggestions

Digital phenotyping research requires careful consideration to maximize data coverage. Solutions include recruiting additional participants^12^ to increase the chance of higher data coverage and study protocols designed to maximize data coverage from enrolled participants. While imputation methods can also help address some issues related to missingness^12^ after data collection, solutions to maximize data coverage are the focus of our recommendations.

To that end, it is important to consider the reasons for low data coverage and design protocols to minimize them. A first consideration is to ensure participants maintain active engagement with the app so that passive data coverage is maximized (see Fig. 3). This may include asking participants to complete surveys in the app, view their results, or any other activity. A second consideration is to work with participants to minimize the times their phone enters low power mode and passive data collection is throttled. Some phones may even stop passive data collection if the phone is off and motionless. A third consideration is to ensure the participants have set the appropriate app permissions and not revoked sensor permission by mistake. Designing protocols and checks around these three considerations is often simple and may result in a large increase in data coverage.

Considering these challenges, we present several practical methods our team has employed that are broadly applicable across any digital phenotyping app or study. These include 1) run-in periods, 2) education and hands-on support, and 3) tools to easily monitor data coverage.

Run-in periods are common in pharmaceutical studies and also have applicability for digital phenotyping. For example, in our second study on college student mental health, we introduced a 3-day run-in period before participants could enter the study. During this time participants completed a 1-2 question survey each day and accelerometer and GPS data collection were monitored. Only participants who were able to complete the surveys each day and achieve a minimum threshold of data collection were able to enter the study. This helped ensure that only participants with a smartphone able to generate sufficient coverage of data and with the ability to actively engage with the app entered the study. In addition, we schedule activities for participants at least once per day to encourage daily interaction with the app. The burden on participants with this method remains minimal as the surveys required less than one minute to complete.

Like any study, participant education and hands-on support can be useful. For example, in our studies using mindLAMP to augment therapy for depression and relapse prediction for schizophrenia we included patient education with Digital Navigator support at the beginning of the study [19]. A Digital Navigator acts as a coach and can reach out to help troubleshoot low data coverage by assessing and addressing common issues (eg incorrectly set sensor permissions). For these studies, the Digital Navigators met virtually with participants at the beginning of the study, walked them through app setup, discussed various reasons that passive data would not collect, and were available if any issues arose.

Another useful approach is to employ tools and procedures that make it easy to monitor passive data quality in near real time. As an example, we have created interactive visualizations on the Data Portal of our researcher dashboard^20^ to allow study staff or clinicians a simple way to track participant data. These graphs are automatically updated each day and contain information about data coverage (as shown in Fig. 7), passive features, and active data. The code to generate these graphs is open source and can be found at https://github.com/BIDMCDigitalPsychiatry/LAMP-cortex. These graphs can be used to help identify participants requiring help with data coverage or even to recognize emerging issues that may arise and threaten data quality across an entire study or cohort. They can also be shared directly with participants, if desired, given that such data sharing itself may prompt more active engagement with the app^21^.

**Fig. 7.**
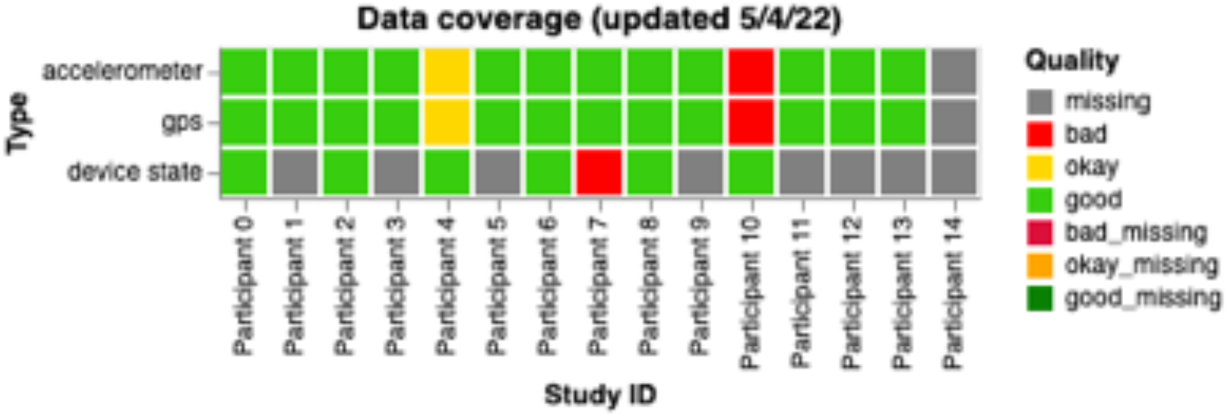
A sample graph for researchers and clinical staff showing data coverage for all participants for a study. Each box is colored based on the particiant’s status: red for bad, yellow for okay, and green for good.

Many prior works have recruited people interested in using one type of device and fewer studies support both Android and Apple smartphones. Like in prior papers [8], we found some differences between Apple and Android groups but did not find a high degree of bias in our studies. Appendix C shows data coverage across phone types and operating system versions. As there was a large variety in our studies, it is difficult to determine whether specific phone types or operating systems should be excluded. Appendix D shows data coverage results for different genders, races, ethnicities, and illness severities in our studies. Although there may be groups, such as certain illness severities, that are unable to achieve an acceptable level of data coverage, the fact that there are few differences between groups in our study seems to suggest otherwise. However, this is a complex topic that requires further investigation.

Combining these methods and focusing on data coverage can lead to positive results. The upward trajectory of data coverage as shown in Fig. 2, especially the increases in Fall 2021 coincide with the implementation of the strategies discussed above. However, the mindLAMP app and smartphone operating systems are constantly changing and improving, so it is difficult to discern whether the data coverage improvements are solely because of these procedures. Regardless, based on our results these proposed solutions should help mitigate the common challenges around data coverage.

## 5 Discussion

In this paper, we discuss the potential, challenges, and solutions towards using smartphone passive data for research or care. Our results shared suggest that while it remains highly feasible to obtain a high degree of passive data coverage, this is likely only possible with multiple concerted active efforts designed to ensure ongoing data coverage.

The results shared in this paper also help explain why many smartphone sensing studies have been challenging to replicate. The likely high degree of variance in raw passive data coverage obtained in different studies suggests that the derived features may not be truly comparable. Given this heterogeneity is related to both the smartphone operating systems as well as the degree of active engagement by each participant with the app, any efforts at replication may be impossible today as few studies report on active app engagement at an individual level. This suggests the clear need for a system of reporting standards that include information on the smartphone operating system, metrics of active engagement, and resulting data coverage metrics if the field seeks to transform passive data-derived features into clinically meaningful insights.

Creating standards will not be simple. Given the nature of this work, stakeholders must include technology manufacturers that control the operating systems, clinicians / researchers seeking to use the data, and patients who must both engage with such systems and ultimately derive value from them. Given the potential scalability of data collection through digital phenotyping methods, such efforts must be global in nature and support privacy and ethical standards across different countries and populations. Creating such standards will not be a simple task but the alternative may be divergent digital phenotyping efforts that are not synergistic, comparable, or impactful.

While such standards are not yet in place, our results can still inform how clinical studies using these methods are both designed and supported. Still even our proposed solutions present challenges. Run-in periods may improve data coverage by excluding those not able to meet minimum thresholds, but today we have little information on which types of people may be excluded as a result. The potential for a high degree of data coverage versus biased results must be considered. Monitoring for bias is thus critical. Digital navigators and data monitoring with dashboards are promising approaches to further increase data coverage, but need to be budgeted into study designs or clinics. Likewise, as the operating systems of smartphones continue to evolve, so must apps generating this data which means that infrastructure budgets need to be reframed from single-time expenses to supporting ongoing maintenance.

There are several limitations of this work. First, the down sampling experiment did not have ground-truth estimates of home time or entropy. However, we still feel that these experiments are valuable in demonstrating how passive data feature estimates can change dramatically if data coverage is low. In addition, we did not have a control group for the data monitoring strategies presented which makes it difficult to determine whether these techniques are the sole cause of the improvements we have seen. However, these solutions offer practical approaches to directly reduce the problems presented.

In summary, the potential of smartphone passive data to generate new research and clinical insights into behavior and illness remains high. But low data coverage of passive data can lead to derived features that are inaccurate and misleading. Active engagement strategies, like planning around low power mode and other operating system changes, can be employed to passive data to increase this coverage and future efforts to establish standards will further increase the reproducibility of this work.

## Data Availability

The code for the app and analysis are available online at https://github.com/BIDMCDigitalPsychiatry

## Funding

Wellcome Trust, National Institute of Mental Health, Sydney Baer Jr. Foundation

## Competing Interests

JT is the cofounder of Precision Mental Wellness, a mental health app company.

## Author Contributions

Both authors contributed equally to this paper

## Data sharing statement

Technical appendix, app, and statistical code available from: https://github.com/BIDMCDigitalPsychiatry

## Acknowledgments

Research assistants who helped run these mindLAMP studies and the people who partook as participants.

## Appendices

**Appendix A:**
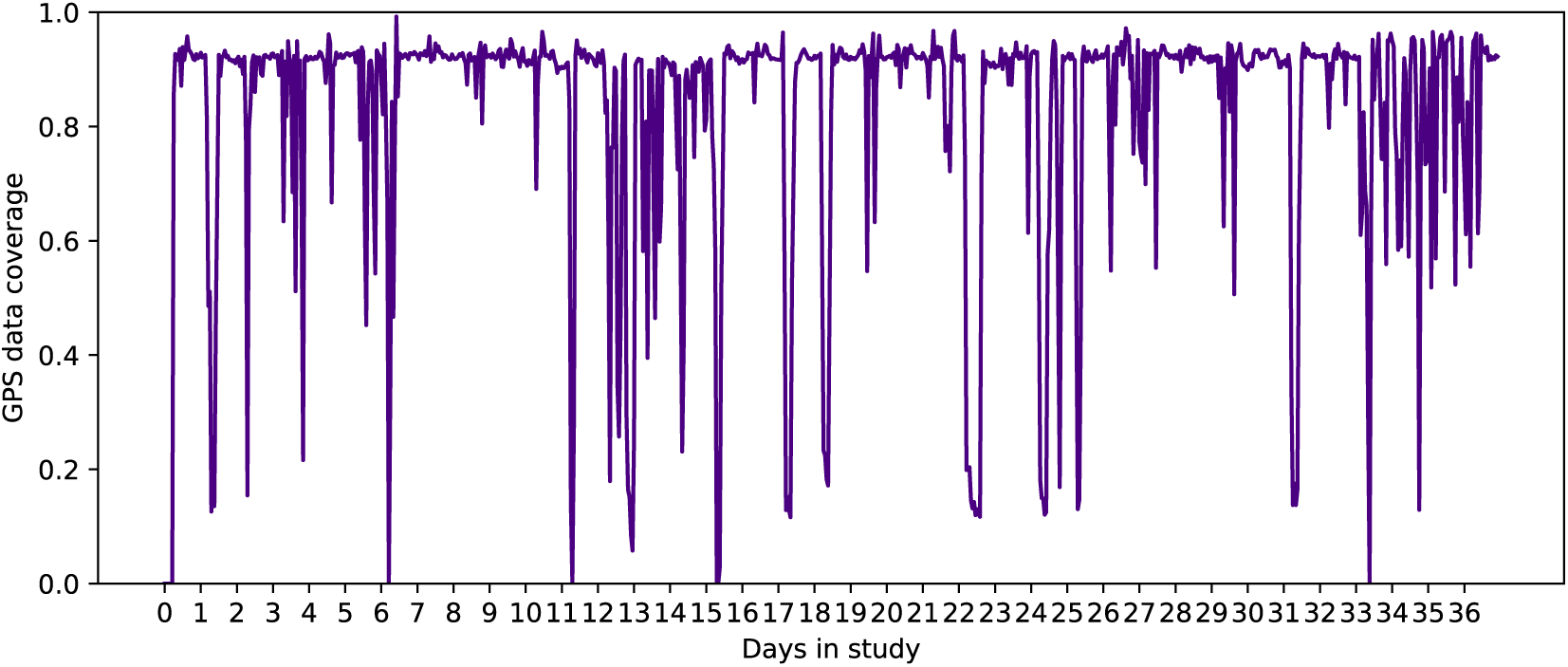
The plot shows the hour-by-hour GPS data coverage for the participant used in the downsampling experiment. The average 1-second data coverage was 83%. The 10-minute data coverage was 98%.

**Appendix B:**
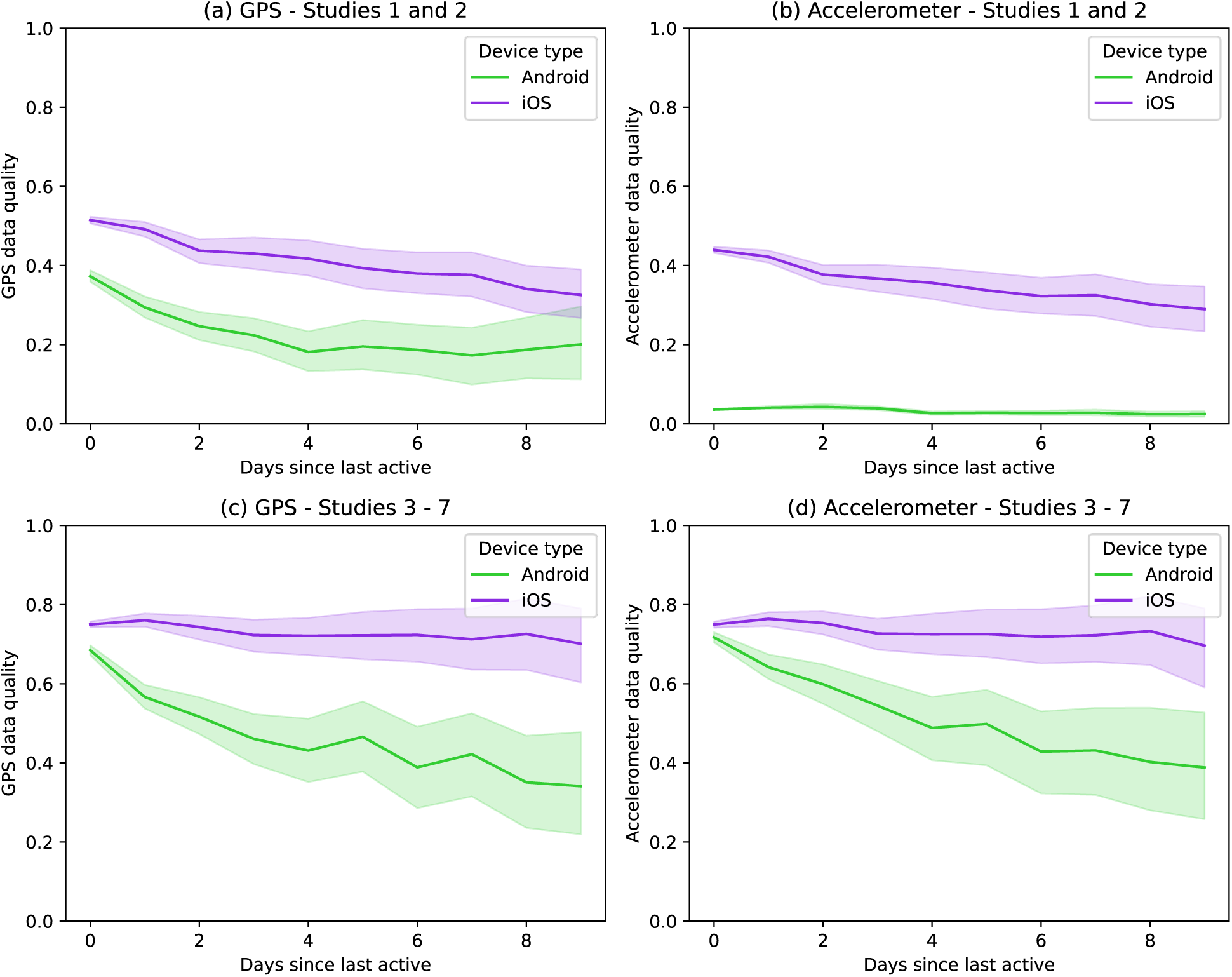
The graphs show the data coverage versus the number of days since the participant last completed an activity in the app. The top row shows (a) GPS and (b) accelerometer coverage for the oldest two studies in this dataset, Study 1 and 2. The bottom row shows (c) GPS and (d) accelerometer coverage for all of the most recent studies (Studies 3-7).

**Appendix C:**
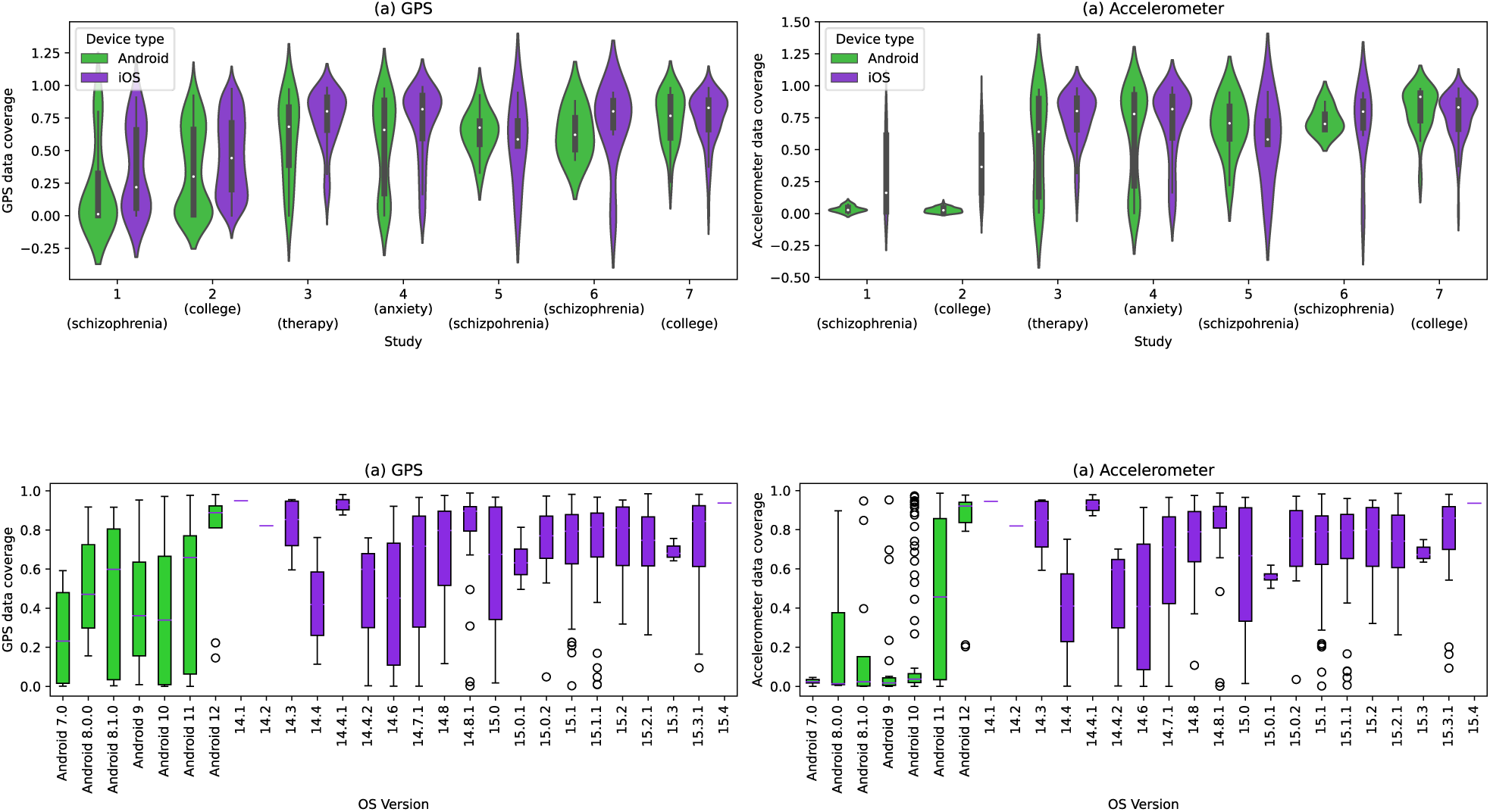
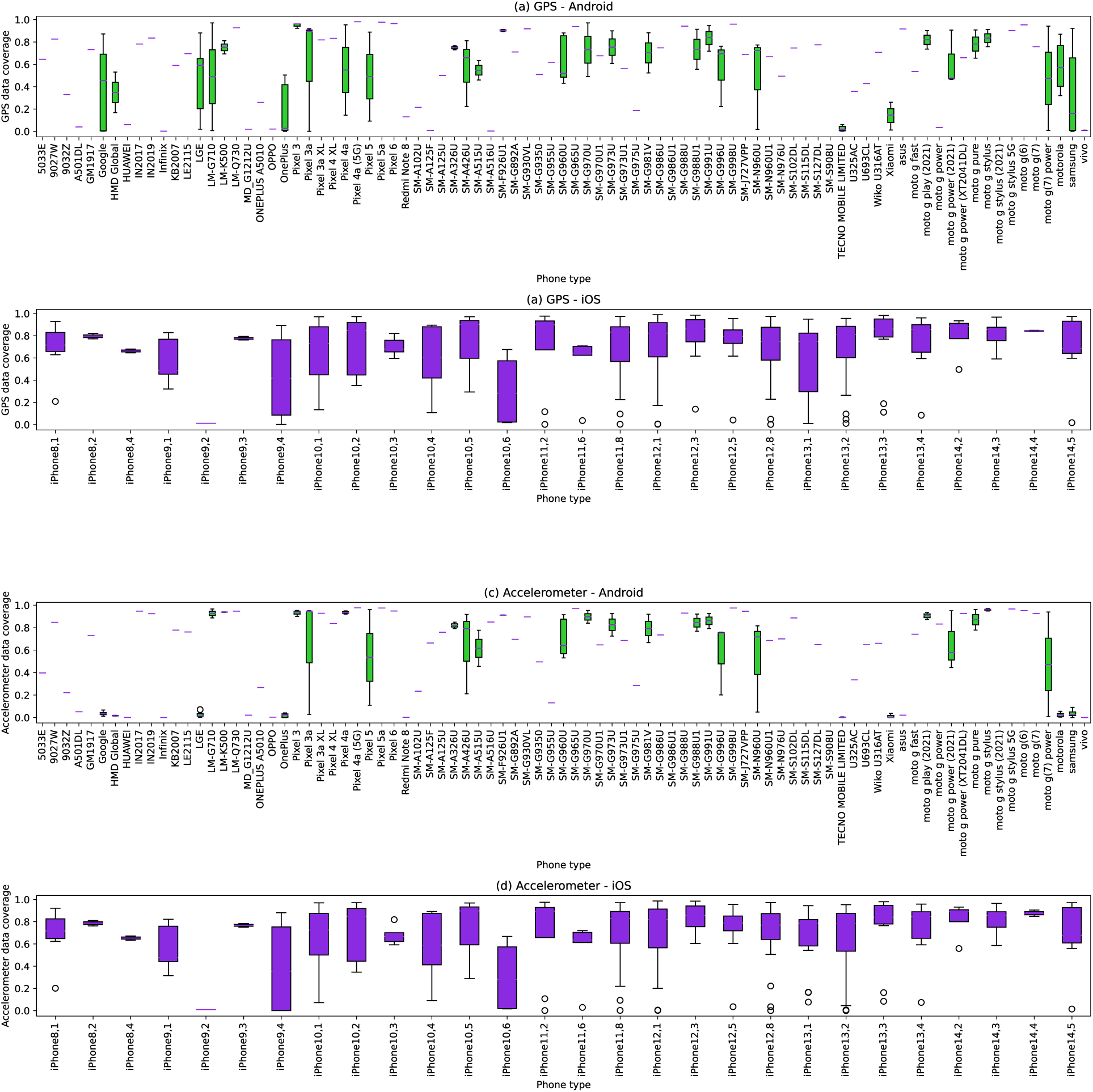
*Appendix C.1:* This plot shows the data coverage across all studies, separate by Android or iOS device.* *Appendix C.2:* These boxplots show the data coverage across different operating system versions across both Android (green) and iOS (purple).* *Appendix C.3:* These boxplots show the data coverage across different phone types across both Android (green) and iOS (purple).* *Device type, operating system version, and phone type was not available for all participants in all studies. All information was available for only 553 participants.

**Appendix D:**
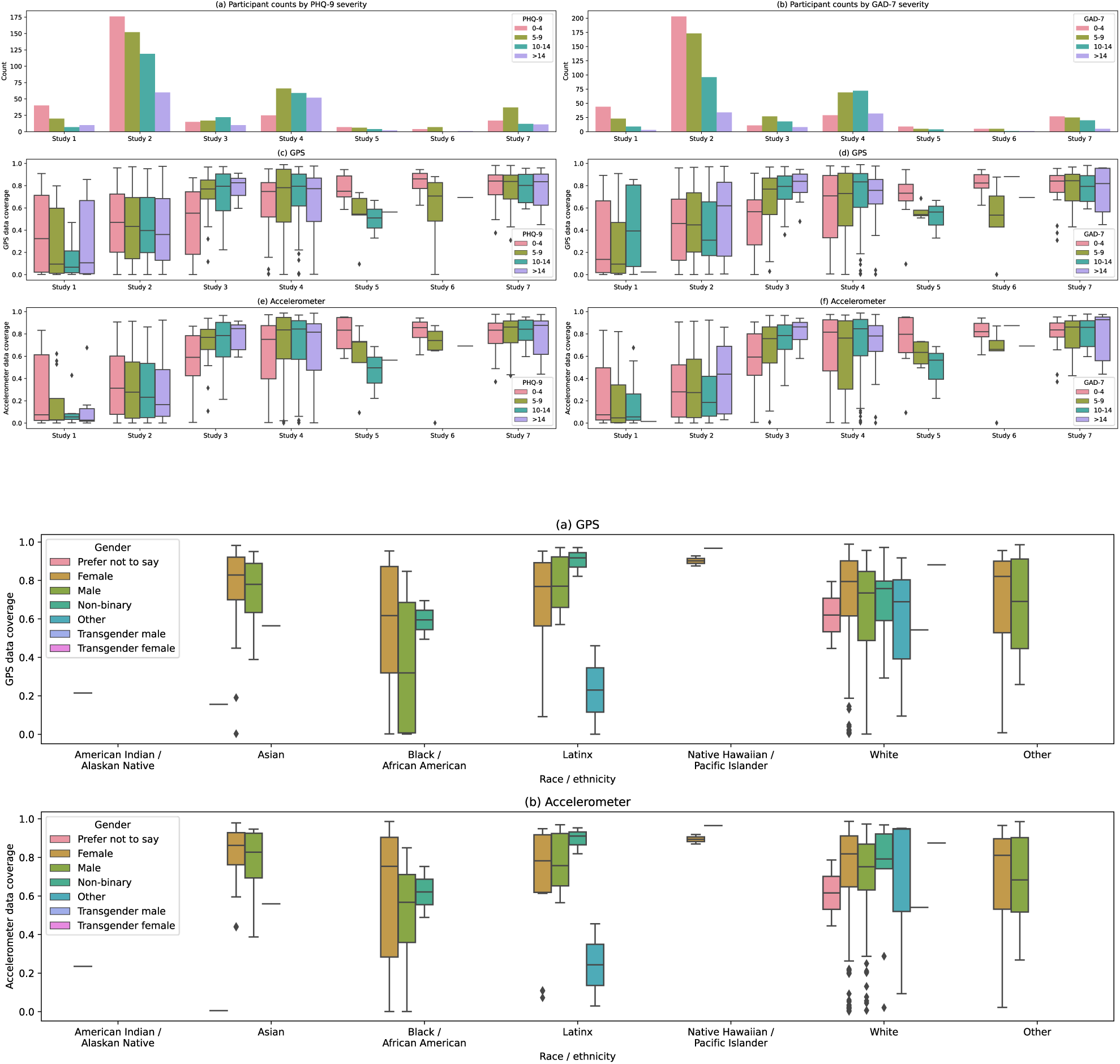
*Appendix D.1:* (a) shows participant counts for each PHQ-9 severity level and (b) shows counts for GAD-7 (groups: <5, 5-9, 9-14, >14). (c) and (e) show boxplots of the data coverage for GPS and accelerometer across the PHQ-9 groups and (d) and (f) show boxplots of the data coverage for GPS and accelerometer across the GAD-7 groups. *Appendix D.2:* Boxplots of (a) GPS and (b) accelerometer data coverage are shown across different race / ethnicity and gender groups. Race / ethnicity and gender information is shown in tables 2a and 2b respectively.

**Table D.3.** Gender information from all studies.

| Study | Total* | Female | Male | Nonbinary | Other** | Transgender Male** | Transgender Female** |
| --- | --- | --- | --- | --- | --- | --- | --- |
| Study 1 (schizophrenia) | 92 | 29 | 45 | 0 | 0 | 0 | 0 |
| Study 2 (college) | 644 | 400 | 225 | 16 | 0 | 0 | 0 |
| Study 3 (therapy) | 92 | 58 | 22 | 3 | 2 | 1 | 0 |
| Study 4 (anxiety) | 220 | 142 | 29 | 6 | 2 | 0 | 0 |
| Study 5 (schizophrenia) | 22 | 10 | 3 | 0 | 2 | 0 | 0 |
| Study 6 (schizophrenia) | 12 | 5 | 5 | 1 | 0 | 0 | 1 |
| Study 7 (college) | 96 | 63 | 28 | 5 | 0 | 0 | 0 |
| Total | 1178 | 707 | 357 | 31 | 6 | 1 | 1 |

\*Some participants chose not to report demographic information which accounts for the discrepancy in counts.
\*\*Only Study 5 and 6 reported if participants were transgender. All other studies only had categories Female, Male, Nonbinary, and Other.

**Table D.4.** Race / ethnicity information from all studies.

| Study | Total* | American Indian / Alaskan Native | Asian | African American | Latinx ** | Native Hawaiian / Pacific Islander | White | Other |
| --- | --- | --- | --- | --- | --- | --- | --- | --- |
| Study 1 (schizophrenia) | 92 | 1 | 2 | 17 | 0 | 0 | 40 | 14 |
| Study 2 (college) | 644 | 2 | 133 | 52 | 117 | 6 | 298 | 34 |
| Study 3 (therapy) | 92 | 0 | 2 | 0 | 8 | 1 | 65 | 9 |
| Study 4 (anxiety) | 220 | 1 | 9 | 9 | 10 | 0 | 145 | 8 |
| Study 5 (schizophrenia) | 22 | 0 | 0 | 4 | 1 | 0 | 9 | 1 |
| Study 6 (schizophrenia) | 12 | 0 | 0 | 1 | 1 | 0 | 9 | 1 |
| Study 7 (college) | 96 | 0 | 30 | 5 | 6 | 1 | 53 | 1 |
| Total | 1178 | 4 | 176 | 88 | 143 | 8 | 619 | 68 |

\*Some participants chose not to report demographic information which accounts for the discrepancy in counts.
\*\*Study 3, 5 and 6 asked for race and Latinx / Non-Latinx separately. In these studies, participants were included in Latinx only if they marked race as “other/multiracial” and Latinx.

## Authors’ contributions

Both authors contributed equally

## Conflict of interest statements

JT is a co-founder of a digital mental health company, Precision Mental Wellness, which is not discussed in this paper

## Role of funding source

None

## Ethics committee approval

All studies were approved by the BIDMC IRB and all participants signed written informed consent.

## Notes

### Competing Interest Statement

The authors have declared no competing interest.

### Funding Statement

This work was supported by the NIMH, Wellcome Trust, and Sydney Baer Jr. Foundation.

### Author Declarations

Beth Israel Deaconess Medical Center gave ethical approval for this work

